# Development and validation of a deep learning algorithm using fundus photographs to predict 10-year risk of ischemic cardiovascular diseases among Chinese population

**DOI:** 10.1101/2021.04.15.21255176

**Authors:** Yanjun Ma, Jianhao Xiong, Yidan Zhu, Zongyuan Ge, Rong Hua, Meng Fu, Chenglong Li, Bin Wang, Li Dong, Xin Zhao, Jili Chen, Ce Rong, Chao He, Yuzhong Chen, Zhaohui Wang, Wenbin Wei, Wuxiang Xie, Yangfeng Wu

## Abstract

**Background:** Ischemic cardiovascular diseases (ICVD) risk predict models are valuable but limited by its requirement for multidimensional medical information including that from blood drawing. A convenient and affordable alternative is in demand.

**Objectives:** To develop and validate a deep learning algorithm to predict 10-year ICVD risk using retinal fundus photographs in Chinese population.

**Methods:** We firstly labeled fundus photographs with natural logarithms of ICVD risk estimated by a previously validated 10-year Chinese ICVD risk prediction model for 390,947 adults randomly selected (95%) from a health checkup dataset. An algorithm using convolutional neural network was then developed to predict the estimated 10-year ICVD risk by fundus images. The algorithm was validated using both internal dataset (the other 5%) and external dataset from an independent source (sample size = 1,309). Adjusted R^2^ and area under the receiver operating characteristic curve (AUC) were used to evaluate the goodness of fit.

**Results:** The adjusted R^2^ between natural logarithms of the predicted and calculated ICVD risks was 0.876 and 0.638 in the internal and external validations, respectively. For detecting ICVD risk ≥ 5% and ≥ 7.5%, the algorithm achieved an AUC of 0.971 (95% CI: 0.967–0.975) and 0.976 (95% CI: 0.973–0.980) in internal validation, and 0.859 (95% CI: 0.822–0.895) and 0.876 (95% CI: 0.816–0.837) in external validation.

**Conclusions:** The deep learning algorithm developed in the study using fundus photographs to predict 10-year ICVD risk in Chinese population had fairly good capability in predicting the risk and may have values to be widely promoted considering its advances in easy use and lower cost. Further studies with long term follow up are warranted.

## Introduction

Ischemic cardiovascular diseases (ICVD), including ischemic stroke and coronary heart disease, were predicted to impose 11.7 million deaths in 2017 globally.^1^ In China, stroke and ischemic heart disease had become the top two causes of death and disability-adjusted life-years in 2017.^2^ ICVD risk prediction model has been used as a valuable tool for primary prevention of ICVD.^3^ A calculated ICVD risk reaching 5% or 7.5% would be identified as reaching borderline or intermediate and initiate additional measurements or interventions.^3^ A sex-specific optimal 10-year ICVD risk prediction model in Chinese adults had been developed and validated.^4^ The model employed a group of parameters: sex, age, systolic blood pressure (SBP), total cholesterol, body mass index (BMI), current smoking or not, and suffering from diabetes or not.^4^ Similar to the Framingham Risk Score in America and the SCORE in Europe, this model required information from questionnaire, invasive blood tests, and physical examinations, which impede its widely use in regular medical check-ups and outpatient clinics.^4-6^

The emergence of non-mydriatic fundus photography offers a prospective and non-invasive approach to handily predict cardiovascular disease. On one hand, acquisition of fundus photographs is of satisfactory convenience.^7^ According to our experience in survey field, a student could take two-sided fundus photographs within one minute after a few hours of training. On the other hand, anatomically and developmentally, the retina is an extension of the brain and a valuable window for observing small vessels in vivo.^8, 9^ Several ICVD risk factors have been associated with corresponding retinal microvascular features.^10, 11^ For example, venular dilatation and retinopathy lesions signal the presence of diabetes or obesity, while arteriolar narrowing was associated with hypertension.^10, 11^ These retinal microvascular abnormalities have also been demonstrated to be associated with increased cardiovascular disease risk.^12, 13^

Deep learning further facilitates and empowers the use of fundus photography. Deep learning algorithms have been developed and validated to screen diabetic retinopathy and multiple abnormal findings in retinal fundus images, with accuracy rivalling that of experts.^14-17^ Poplin et al. show that deep learning can be used to discriminate a group of cardiovascular risk factors from retinal fundus images, including sex, age, ethnicity, BMI, SBP, HbA1c, and current smoking.^18^ However, studies examining the association between fundus images and ICVD 10-year risk are sparse.

Thus, the present study aimed to develop and validate a deep learning algorithm to predict 10-year ICVD risk using retinal fundus photographs in Chinese population.

## Methods

### Study design

This was a cross-sectional study. We developed an algorithm using convolutional neural network from a medical checkup data. The algorithm can be used to predict a calculated 10-year ICVD risk using retinal fundus image. Then the algorithm was validated using internal dataset from the same checkup data and an independent dataset.

### Datasets

For algorithm development, a dataset of 411,518 individuals from Tongren Hospital in Beijing, Shibei Hospital in Shanghai, and conventional medical check-up population in 19 province-level administrative regions of China during Sep 2018 to Dec 2019 was used. The dataset contained fundus images of both eyes and medical information, including sex, age, SBP, total cholesterol, BMI, and suffering from diabetes or not. The individuals in the check-up population were simply randomized into a development dataset (95%) and an internal validation dataset (5%). The use of this dataset in this study was approved by Tongren Hospital Institutional Review Board, Shibei Hospital Institutional Review Board, and iKang Healthcare Group Institutional Review Board with a waiver of informed consent.

The performance of the algorithm was further validated using the data of the Beijing Research on Ageing and VEssel (BRAVE). The BRAVE draw participants who aged 45 to 75 years and lived in Shijingshan District, Beijing in 2019. Participants with fundus images, complete information of all the parameters for 10-year ICVD risk calculation, and without ICVD history were included. This community-based cohort was approved by the Peking University School Institutional Review Board and conformed to the Declaration of Helsinki. Written informed consent was obtained from each participant.

A variety of cameras were used in obtaining fundus images, including Canon CR1/CR2 and Crystalvue FundusVue/TonoVue in the check-up dataset and Centervue DRS in the BRAVE. All images were captured using 45° fields of view. The Cox function for 10-year ICVD risk prediction in Chinese adults proposed by Wu et al. was used in both datasets, which involved seven parameters including sex, age, SBP, total cholesterol, BMI, current smoking or not, and suffering from diabetes or not.^4^ The risk calculation approach used by Wu et al. was rigorously followed in the BRAVE, but not in the check-up dataset for missing data^4^. Sex and age were self-reported in both datasets. Current smoke status was self-reported in the BRAVE but not available in the check-up dataset. The smoking status in check-up dataset was imputed according to age. Total cholesterol was measured using fasting blood samples in both datasets. Diabetes was defined as self-reported doctor-diagnosed history or a fasting serum glucose level ≥ 126 mg/dL (7.0 mmol/L) in the BRAVE, but only according self-reported history in the check-up dataset for missing fasting serum glucose data. In the BRAVE, SBP was took for three times with one-minute intervals, the mean of which was used. But only one SBP measurement was performed in the check-up dataset.

### Development of the algorithm

The development dataset was divided into two parts by image: a training dataset and a tuning dataset. The tuning dataset was used for selecting model with best performance during training. As the Cox function for 10-year ICVD risk calculation is an exponential function that may have surge of output risk due to small input variation, which could earn much of neural network optimization attention. Eventually, this may ruin the model parameters. Hence, we use natural logarithm of ICVD risk to avoid the surge. The ICVD label y_ICVD_ for training and testing of the network is given as y_ICVD_ = ln(risk_ICVD_ × 100), where risk_ICVD_ is the 10-year ICVD risk obtained from Cox regression function. The detailed calculation process can be found in eMethods and eTable 1 the Supplemental material. This logarithm operation has no loss of generality on the prediction capability of Cox function.

**Table 1.** Characteristics of individuals in development, internal validation, and external validation datasets

| Characteristics | Development dataset |  | Internal validation dataset |  | External validation dataset |  |
| --- | --- | --- | --- | --- | --- | --- |
|  | Men | Women | Men | Women | Men | Women |
| No. of images | 426,215 | 372,651 | 23,068 | 19,982 | 940 | 1,674 |
| No. of participants | 208,398 | 182,549 | 11,001 | 9,570 | 470 | 837 |
| Age | 42.1±13.3 | 41.7±13.4 | 42.2±13.4 | 41.6±13.3 | 60.4±7.1 | 58.8±7.2 |
| SBP/mm Hg | 124.2±16.3 | 116.1±17.7 | 124.4±16.2 | 116.1±17.7 | 137.3±16.3 | 131.6±17.7 |
| total cholesterol | 4.9±0.9 | 4.9±0.9 | 4.9±0.9 | 4.9±1.0 | 4.8±0.9 | 5.0±0.9 |
| BMI/ (Kg/m<sup>2</sup>) | 25.0±3.4 | 22.7±3.4 | 24.9±3.5 | 22.8±3.4 | 26.1±3.2 | 25.8±3.5 |
| Currently Smoking* | - |  | - |  | 246 (52.3) | 26 (3.1) |
| Diabetes | 13,202 (3.1) | 6,566 (1.8) | 757 (3.3) | 340 (1.7) | 121 (25.7) | 172 (20.6) |

Data are presented as are presented as mean ± SD or n (%).
\* Smoking status was not available in development or internal validation datasets.

This work used deep learning neural network to learn ICVD prediction from fundus image. Inception-Resnet-v2 was used as the backbone of the deep learning architecture with input size of 299×299.^19^ For the input, the backbone returned an output with a dimension of 8×8×1536 which was forwarded to 8×8 size average pooling layer and then been flatten into a 1×1536 dimension vector. A fully connected layer of a unit size of 32 was subsequently used, which is connected to a dense unite to return ICVD prediction. The mean absolute error was incorporated as the loss function for the prediction. Adam method was adopted for network parameter optimization (batch size of 64) with an initial learning rate at 0.001.^20^ The Adam algorithm tuned the parameters for ICVD prediction optimization according to the MAE loss calculated from the predicted value and y_ICVD_.

In the training of the neural network, data augmentation methods included random cropping, random rotation (±30°) and random horizontal flipping. A preprocess is applied to the input to map pixel values into a given distributions in both training and testing of the model.^21, 22^ The model is trained on Keras platform v2.2.2 and Python scikit-learn package 0.22.2 on a computer equipped with Nvidia GeForce GTX 1080Ti GPU ×2.^23^

### Validation of the algorithm

The performance of the algorithm was evaluated using coefficient of determination (R^2^) and area under the receiver operating characteristic curve (AUC). The trained neural network was used to yield predicted value of y_ICVD_ from fundus images each participant in the internal validation dataset and the BRAVE. The mean y_ICVD_ of both eyes was used to deduce the predicted risk_ICVD_. At the same time, calculated y_ICVD_ and risk_ICVD_ were generated from parameters, including sex, age, SBP, total cholesterol, BMI, current smoking or not, and suffering from diabetes or not.^4^ The R^2^ between the predicted y_ICVD_ and calculated y_ICVD_ was obtained. Since 5% and 7.5% was usually regarded as the thresholds for 10-year ICVD risk classification^3^, the AUC of the algorithm were computed for screening individuals with calculated risk_ICVD_ ≥ 7.5% with 95% CI by the pROC package version 1.16.2.^24^

## Results

The characteristics of individuals in the development dataset, internal validation dataset, and the BRAVE are summarized in Table 1. The development and internal validation datasets shared almost the same characteristics. They included 390,947 (mean aged 41.9 ± 13.4 years, men: 53.3%) and 20,571 (mean aged 41.9 ± 13.4 years, men: 53.5%) participants, respectively (Figure 1a). The characteristics of individuals in the training and tuning datasets were presented in eTable 2 in the Supplemental material. Among 1,554 participants attending the baseline survey in the BRAVE, 1,305 participants (mean aged 59.4 ± 7.2 years, women: 64.0%) had fundus photos of both eyes, complete data of the parameters for calculating 10-year ICVD risk, and no ICVD history (Figure 1b).

**Figure 1.**
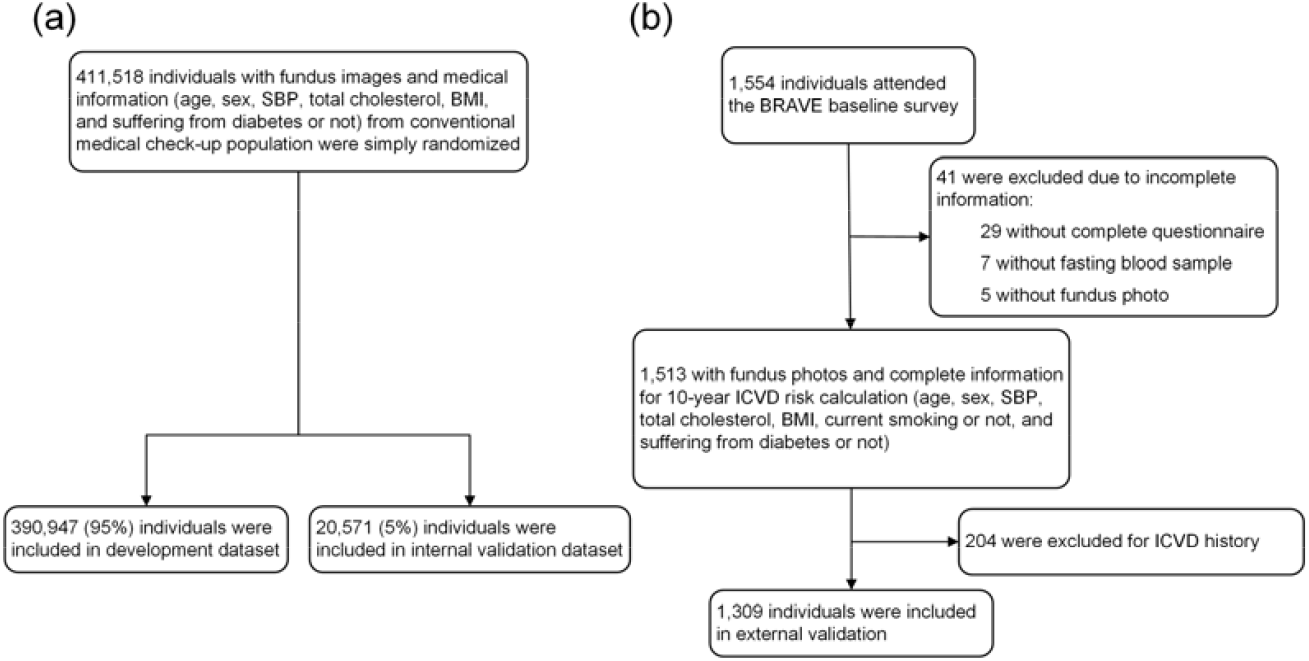
Flow chart of participants selection in development, internal validation, and external validation datasets.

The adjusted R^2^ between natural logarithms of the predicted and calculated ICVD risks, or the predicted and true y_ICVD_, was up to 0.876 in the internal validation dataset and 0.638 in the BRAVE (Figure 2). Respectively, 774 (3.8%) individuals in the internal validation dataset and 68 (5.2%) in the BRAVE had calculated 10-year ICVD risk ≥ 5%, indicating borderline or higher ICVD risk. At the same time, 496 (2.4%) in the internal validation dataset and 27 (2.1%) in the BRAVE had calculated 10-year ICVD risk ≥ 7.5%, indicating intermediate or higher ICVD risk. For detecting calculated ICVD risk ≥ 5% and ≥ 7.5% in the internal validation, the algorithm achieved an AUC of 0.971 (95% CI: 0.967–0.975) and 0.976 (95% CI: 0.973–0.980), respectively (Figure 3); that in the BRAVE were 0.859 (95% CI: 0.822–0.895) and 0.876 (95% CI: 0.816–0.837), respectively (Figure 4). The maximum Youden index were 0.841, 0.857, 0.609, and 0.641, respectively on the four receiver operating characteristic curves (Figures 3 and 4).

**Figure 2.**
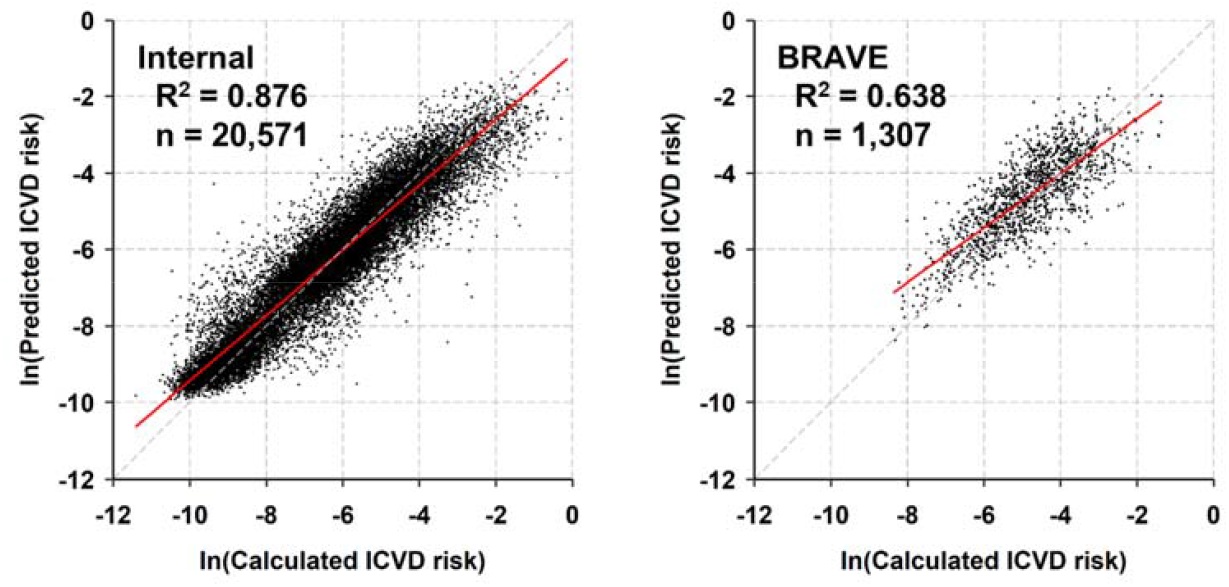
Prediction of natural logarithm of 10-year ICVD risk (or y_ICVD_)

**Figure 3.**
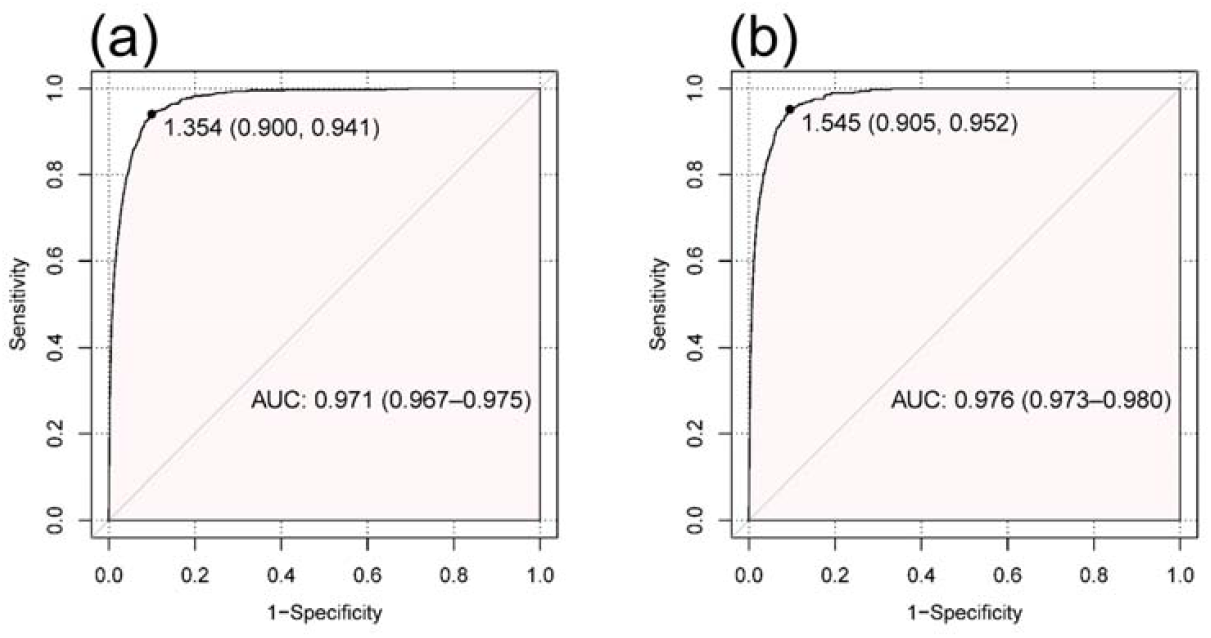
Algorithm for screening individuals with calculated ICVD risk ≥ 5% (a) or ≥ 7.5% (b) in the internal validation dataset

**Figure 4.**
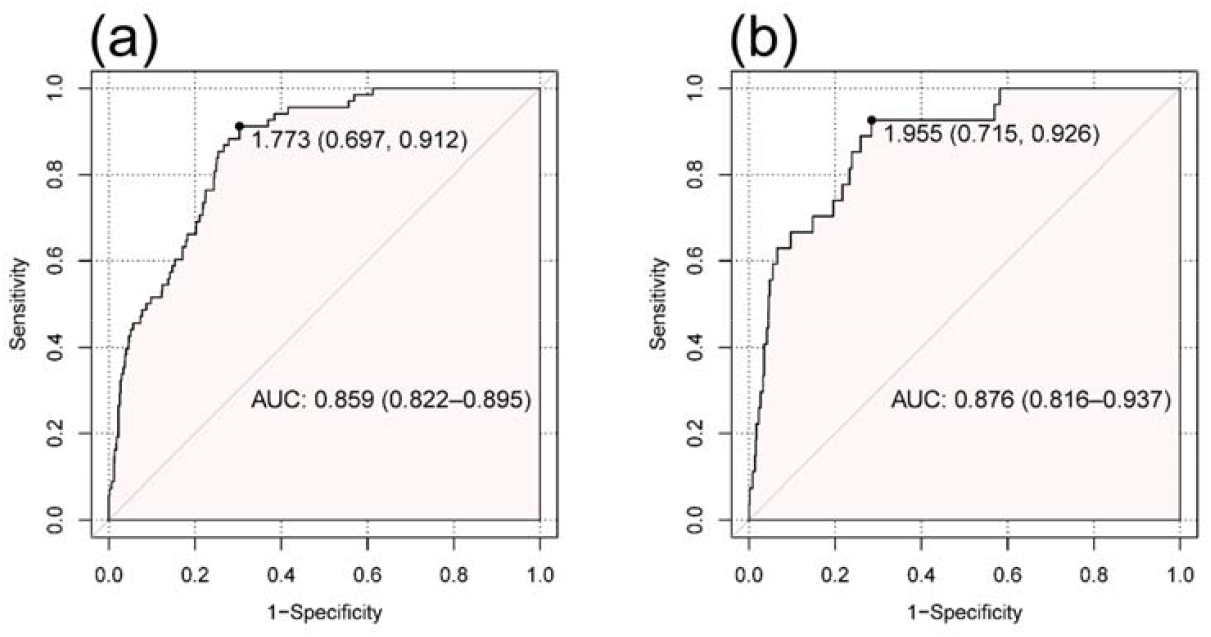
Algorithm for screening individuals with calculated ICVD risk ≥ 5% (a) or ≥ 7.5% (b) in BRAVE

The points on line indicate the threshold of ICVD in percent (specificity, sensitivity) for maximum Youden index.

The points on line indicate the threshold of ICVD in percent (specificity, sensitivity) for maximum Youden index.

## Discussion

This study developed a deep learning algorithm to predict 10-year ICVD risk by fundus photos and validated it in an internal dataset and an independent dataset. The algorithm performed well in screening for individuals with borderline/intermediate or higher ICVD risk, the AUCs of which were over 0.9 in the internal validation and over 0.8 in the external validation. This algorithm is promising to become a convenient approach for ICVD risk screening in clinic or community-based check-ups.

The finds are in consistence with previous studies.^18, 25^ Recently, a study of 155,449 participants demonstrated that normal aging and vascular pathologic changes presented different features in fundus, which could be noticed by convolutional neural network.^25^ Poplin et al. from Google had developed and validated an algorithm to predict a group a cardiovascular risk factors from fundus images among Caucasian and Hispanic, including age, gender, smoking status and SBP.^18^ The algorithms developed by Poplin et al. even could be used to predict major cardiac events in 5 years(AUC = 0.70, 95% CI: 0.65 – 0.74).^18^ While the lipid panels and diabetes diagnosis were unavailable in study by Poplin et al..^18^ What’s more, there are different risk factor patterns and cardiovascular disease profiles between Caucasian and Chinese population.^1, 4^ For example, the leading cause of death was CHD globally and stroke in China.^1, 4^ Thus the algorithm developed in the present study is relevant and would facilitate the application of deep learning in clinical practice in China.

The demographic characteristics were significantly different in the development dataset and the BRAVE. Participants in the validation dataset was older, contained more women in proportion. Given those differences, the algorithm still performed well in screening borderline/intermediate or higher ICVD risk, which indicated the robustness of this algorithm. Hence, the algorithm could be used as an alternative to traditional ICVD prediction model for Chinese population. Although the traditional model by Wu et al. had been validated and extensively used in researches, it had not been widely used in clinics or regular medical checkups.^4, 26-28^ The probable cause included its requirement for multidimensional medical information and complex calculation. In addition, limited primary care physician consultation length of only 2.0 minutes might made it more infeasible to apply traditional ICVD prediction model.^29, 30^ In comparison, the average consultation length was over 20 minutes in USA.^30^ As an alternative to traditional model, this approach to assess ICVD risk using fundus photos and deep learning algorithm is much more speedy and affordable. Taking non-mydriatic fundus of both sides only costs less than one minute and six RMB (less than one USD) per person in Beijing.^31^ Thus, such an alternative would be significant for clinical practice and public health, especially in populations with insufficient healthcare resources.

The findings should be interpreted with some caution limitations of the study. First, the 10-year ICVD risk was calculated according to parameter using cross-sectional data rather than drew from the real ICVD events in longitudinal studies, which limited the reliability of the algorithm. Second, data of current smoking status was absent in the development dataset. In this regard, age and sex was used to predicte current smoking status in development and internal datasets. While the findings from the external validation was not interfered by this data missing.

## Conclusions

This study demonstrated that application of deep learning algorithm to fundus photographs along can be used to predict 10-year ICVD risk in Chinese population. As an expedient alternative of traditional models, the algorithm performed well in screening for individuals with borderline/intermediate or higher ICVD risk. The effectiveness of applying this algorithm in clinic or regular physical checkups are needed before its wide application. And further deep learning researches on the association between fundus images and subsequent ICVD incidence of prospective cohort design are warranted.

## Data Availability Statement

The datasets can also be obtained on request.

## Data Availability

The data can also be obtained on request.

## Acknowledgments

We appreciate efforts made by the original data creators and their contributions for access of data.

## Sources of Funding

The present study was supported by the National Natural Science Foundation of China (project no. 81974489, 81974490).

## Disclosures

None.

## Notes

### Competing Interest Statement

The authors have declared no competing interest.

### Author Declarations

%). The use of this dataset in this study was approved by Tongren Hospital Institutional Review Board, Shibei Hospital Institutional Review Board, and iKang Healthcare Group Institutional Review Board with a waiver of informed consent.

