## Supplemental for "Development and validation of a deep learning algorithm using fundus photographs to predict 10-year risk of ischemic cardiovascular diseases among Chinese population"

**eMethods**

The 10-year ICVD risk was calculated from sex, age, systolic blood pressure (SBP), total cholesterol, body mass index (BMI), current smoking or not, and suffering from diabetes or not. The risk was calculated using the equation: $Risk=1-S^{e^{X-mean（X）}}$, where X was for the sum of “Coefficient × Value” column in eTable 1, S for baseline survival function at 10 years (eTable 1).

**eTable 1. Characteristics of individuals in training and tuning datasets**

| **Characteristics** | **Men** | | |  | **Women** | | |
| --- | --- | --- | --- | --- | --- | --- | --- |
|  | **β** | **Individual example**  **Value*** | **Coefficient × Value*** |  | **β** | **Individual example**  **value†** | **Coefficient × Value†** |
| Age, 5 year | 0.3277 | 13 | 4.2601 |  | 0.4641 | 18 | 8.3538 |
| SBP, 20 mm Hg | 0.6711 | 6.85 | 4.5970 |  | 0.6090 | 6.70 | 4.0803 |
| Total cholesterol, 1 mmol/L | 0.1367 | 5.60 | 0.7655 |  | 0.1479 | 4.75 | 0.7025 |
| BMI, 3 kg/m2 | 0.1360 | 9.1 | 1.2376 |  | 0.1593 | 9.0 | 1.4337 |
| Current smoker, yes/no | 0.6894 | 1 | 0.6894 |  | 0.4457 | 0 | 0.0000 |
| Diabetes, yes/no | 0.1195 | 0 | 0.0000 |  | 0.9946 | 1 | 0.9946 |
| Individual sum (or X) | - | - | 11.5497 |  | - | - | 15.5649 |
| Mean (X)‡ | - | - | 10.8006 |  | - | - | 11.7962 |
| Baseline survival function at 10 years (or S) | - | - | 0.9835 |  | - | - | 0.9948 |
| Calculated 10-year ICVD risk, percent | - | - | 3.4578 |  | - | - | 20.2179 |

*A male example: 65 years of age with mean SBP 137.0 mm Hg, total cholesterol 5.60 mmol/L, BMI 27.3 kg/m2, smoker, and without diabetes.

†A female example: 59 years of age with mean SBP 134.0 mm Hg, total cholesterol 4.75 mmol/L, BMI 27.3 kg/m2, nonsmoker, and with diabetes.

‡The mean values of X in the BRAVE are used.

**eTable 2. Characteristics of individuals in training and tuning datasets**

| **Characteristics** | **Training dataset** | |  | **Tuning dataset** | |
| --- | --- | --- | --- | --- | --- |
|  | **Men** | **Women** |  | **Men** | **Women** |
| No. of images | 404,250 | 353,464 |  | 21,965 | 19,187 |
| No. of participants | 207,844 | 182,113 |  | 21,310 | 18,687 |
| Age | 42.1±13.3 | 41.7±13.4 |  | 42.1±13.4 | 41.6±13.4 |
| SBP/mm Hg | 124.2±16.3 | 116.1±17.7 |  | 124.2±16.3 | 116.1±17.6 |
| total cholesterol | 4.9±0.9 | 4.9±0.9 |  | 4.9±0.9 | 4.9±0.9 |
| BMI/ (Kg/m2) | 25.0±3.4 | 22.7±3.4 |  | 25.0±3.4 | 22.7±3.4 |
| Diabetes | 6,262 (3.1) | 3,099 (1.8) |  | 679 (3.1) | 369 (1.9) |

Data are presented as are presented as mean ± SD or n (%).

* Smoking status was not available in development or internal validation datasets.
